# Multimodal Prehabilitation in People Awaiting Acute Inpatient Cardiac Surgery (PreP-ACe): A single-arm feasibility study

**DOI:** 10.64898/2026.11.17.26368355

**Authors:** Sarah Raut, Aaron Hales, David Yates, Gerard Danjoux, Lee Ingle

## Abstract

**Introduction:** The average waiting time for urgent coronary artery bypass graft (CABG) is 13 days in England and Wales and 27 days in Northern Ireland; significantly longer than the National Institute for Cardiovascular Outcomes Research (NICOR) recommendation of 7 days. Inpatients are at risk of Hospital-Associated Deconditioning (HAD) which impacts cognitive and physical function, increases mortality and healthcare cost. Prehabilitation has been shown to improve physical function and reduce complications in the elective population. Prehabilitation has never been attempted in acute inpatients waiting for cardiac surgery.

**Methods:** Participants were recruited between April 2024 and February 2025. Once consented, the participants were provided with psychoeducation booklet and underwent assessments; 6 minute walk test (6MWT), hand grip test (HGT), spirometry, EQ-5D-5L and cardiac anxiety questionnaire (CAQ) on day 0, 7 and 14. The physiotherapy team assessed and prescribed personalised exercise. Information regarding their in-hospital stay were collected. Our primary outcome was determining the feasibility of prehabilitation. The secondary outcome was to identify any adverse events related to the intervention and changes in the assessment results.

**Results:** 17 participants were recruited resulting in a recruitment rate of 1.5 participants per month. 1 participant withdrew from the study. 7 participants were seen by physiotherapist and participated in the exercise program. The adherence to the exercise program was 100% with tolerability of >80%. No adverse events related to exercise recorded. 11 participants remained in the study for 7 days or more.

**Discussion:** Prehabilitation is a novel concept of preparing inpatients for acute cardiac surgery. There are several challenges in determining the feasibility of prehabilitation in this population. These include identifying the best time to recruit, optimal level of exercise, funding (staffing to execute the intervention) and clinician reception towards the intervention. However, prehabilitation was well accepted, tolerated and adhered to by participants.

## Introduction

Prehabilitation is the concept of preparing a patient for an upcoming treatment with the aim of achieving a better post-treatment outcome. There is no single protocol on how to run a prehabilitation program; however, it should include nutrition, exercise and psychological interventions.^(1)^ Exercise has been the main focus of prehabilitation but there is increased awareness that nutrition^(2)^ and psychological^(3, 4)^ factors impact patient recovery. The exercise could include either a single component or combination of respiratory^(5, 6)^, flexibility^(7)^, strength/ resistance^(5)^ and aerobic^(5, 7, 8)^ exercises. Several studies on elective patients waiting for cardiac surgery have shown that exercise in this group is not only safe^(5-7, 9-11)^ but potentially beneficial to their recovery^(10-13)^ and improves their functional capacity.^(5, 7, 9, 11, 13)^

The national target is for urgent coronary artery bypass graft (CABG) to be performed within 7 days of surgery. Despite the reduction in cardiac surgical procedures, the average waiting time in England and Wales is 13 days and 27 days in Northern Ireland.^(14)^ Inpatients are often discouraged from leaving their bedside which leads to deconditioning.^(15)^ Hospital-Associated Deconditioning (HAD) not only negatively impacts cognitive and physical functioning but also leads to increased mortality.^(16)^ Reduced function due to HAD leads to longer hospital stay which translates to an increase in healthcare cost.^(16)^

Prehabilitation has not been done in patients awaiting acute inpatient cardiac surgery. Reasons for this include concerns for safety, feasibility and efficacy given their acute admission as well as short waiting time before surgery. However, prehabilitation as short as 2 weeks has been shown to reduce post operative complications^(10)^, improve functional capacity^(7)^ and quality of life^(7)^. Lopez-Hernandez et al found no association between duration of program or number of training session and positive response to prehabilitation. ^(11)^

The combination of longer waiting time for acute cardiac surgery, risk of deconditioning and evidence of benefit of prehabilitation as short as 2 weeks led us to investigate the feasibility of prehabilitation in acute inpatients waiting for cardiac surgery.

## Methods

### Trial design

This was a single-centre, single-arm feasibility study done in Castle Hill Hospital (CHH), UK. The study aimed to recruit 20 participants over a one year period.

### Ethics and registration

The PreP-ACe trial was ethically approved by the Health Research Authority and Health and Care Research Wales (Yorkshire & the Humber-Bradford Leeds Research Ethics Committee: REC reference 23/YH/0255) on the 8th November 2023. The trial sponsor was the University of Hull. The trial was registered at ClinicalTrials.gov (identifier: NCT06275100). Written informed consent was obtained from all participants. The protocol for this trial was previously published. The trial is reported according to CONSORT guidelines.^(17)^

### Participants

Patients admitted to CHH with cardiac disease between April 2024 and February 2025 were screened for eligibility by the main researcher (first author). All adult patients with surgical pattern of disease were eligible. Patients who were medically unstable, had cognitive and physical impairment that could impede ability to participate in the program, unable to communicate in English or had a date of surgery ≤ 7 days were excluded.

Medically unfit or unstable patients were those with conditions such as chest pain at rest, uncontrolled arrhythmias, and untreated acute heart failure. Identification of potential participants was performed in two stages; the first based on reviewing patient’s medical notes and discussion with the medical team; the second stage involved in person assessment by the researcher to confirm their eligibility. Eligible patients were given participant information sheet. The researcher then returns to enquire about their interest in participating, answer questions and obtain written informed consent.

### Assessments/ Outcomes

The primary outcome is the feasibility of prehabilitation in this group of patients. This includes recruitment, retention, completion, tolerability and adherence. Once consented, baseline assessments were done. The secondary outcome is to identify any adverse events related to the interventions and changes in the assessment results. The baseline assessments were hand grip test (HGT), 6 minutes walk test (6MWT), spirometry, EQ-5D-5L, and Cardiac Anxiety Questionnaire (CAQ). The assessments will be repeated on day 7 and 14 of the study unless the participant had their surgery or were discharged.

Participants who had chest pain at rest did not perform 6MWT.

### Exercise

The exercise consisted of three components; aerobic, strength and flexibility and respiratory muscle training. The aerobic exercise was of moderate intensity, performed under direct supervision and monitoring of the physiotherapist for a maximum of 15 minutes. The exercise was terminated before the 15 minutes if the participants felt unable to continue, exhibited cardiac symptoms or if they achieved moderate intensity which corresponds to 40-70% heart rate reserve (HRR) and Borg Rating of Perceived Exertion (RPE) of 10-14. In addition to this, participants were taught and prescribed self-directed strength, flexibility and respiratory muscle training. Full details of the exercise protocol has been previously published in Plos One.^(18)^

### Psychological support

A psychoeducation booklet was developed with the input of a hospital clinical psychologist. The booklet contained information to help them to rationalise feelings that an inpatient may have, suggest coping strategies and signpost them if they require further assistance.

The main researcher followed the participant’s progress after the prehabilitation to collect data on complications, length of stay and mortality.

### Adverse events

Adverse events were defined as untoward medical occurrences that could lead to mortality or morbidity during or within 30 minutes after the intervention. Adverse events during the exercise were recorded by the physiotherapist. Participants were encouraged to report any cardiovascular symptoms such as chest pain and palpitations to the medical team; this would be recorded in their notes. In addition, SR directly enquires about their symptoms prior to the assessment.

In our previously published protocol, a stop-go criteria was included for the aerobic exercise component of the study. Three criteria included were adherence, tolerability and serious adverse events attributed to the program. Based on the stated criteria, adherence or tolerability of 50-69% or 2-3 serious adverse events would trigger a modification of protocol whereas adherence or tolerability of <50% or more that 3 adverse events attributed to the program would trigger a stop in the protocol.^(18)^

### Statistical methods

Recruitment data was presented as counts over two flow charts. Quantitative data were presented as counts, percentages, means and standard deviation where appropriate in corresponding tables. Descriptive data analysis was performed on Stata MP 19.5. No hypothesis test was performed due to the small data set.

## Results

### Recruitment and completion

482 patients admitted to cardiology ward were screened between 4/4/24 to 28/2/25.(Flow chart 1) The study was terminated early due to staffing issues which impacted the delivery of the program. The majority (423) of these patients were medical patients that did not need surgery. This leaves 59 (12.2%) patients who were possibly for surgery.

These patients were then screened for eligibility for the study by going through the medical notes and speaking to the staff. 21/59 (35.6%) patients did not fit the initial screening criteria as they had a surgery date within 7 days (10 patients), another 10 were medically unfit and 1 was unable to communicate in English; which leaves 38 (64.4%) patients to be considered. 13 (34.2%) patients were unsuitable after further assessment and discussion with the team; hence, they were not approached. 25 patients were approached by the researcher, these were patients that had disease that are potentially operable. Some were awaiting further investigations, weekly multidisciplinary team meeting, surgical review or having alteration in their medication. 8(32%) declined and 17(68%) were recruited. (Flow chart 1) Of the 8 patients that declined to participate, 4 were initially interested but were discouraged by their primary clinical team from participating; they were instead told to have bed rest. 17 participants were recruited over an 11 month period giving a recruitment rate of 1.5 participants per month.

### Summary of participants

All 17 patients completed their initial baseline assessment (test 1) giving a completion rate of 100%. (Flow chart 2) This consisted of HGT, 6MWT, spirometry, EQ-5D-5L and CAQ. In the following days after being recruited, 5 participants were discharged; 2 after percutaneous intervention (PCI), 2 for ‘outpatient surgery’ and 1 for outpatient cardiac MRI. 1 participant declined physiotherapy exercise and a repeat assessment on day 7. The participant was asked if he remembered consenting; which he did, hence we clarified if he would still like to participate in the study. The participant expressed that they ‘couldn’t be bothered with effort’ and ‘just wanted to have the surgery’. As such they were withdrawn from the study. The study had a retention rate of 94.1%.

11 participants should have a second round of assessments (test 2) on day 7 but only 8 (72.7%) participants completed the second test.(Flow chart 2) 2 of the participants did not complete their assessment as SR had a family emergency. 1 participant declined their second assessment as they had a stressful day after medical investigations and speaking to the surgeon. However, they wanted to remain in the study and have otherwise been engaging in the program. 2 participants subsequently had their surgery between day 8 and 14.

6 participants were left in the study for test 3.(Flow chart 2) 1 participant was isolated for MRSA. 5 (83.3%) participants had their final assessment-test 3. 3 (71.4%) were fully completed as 2 participants informed SR that they have been having chest pain at rest.

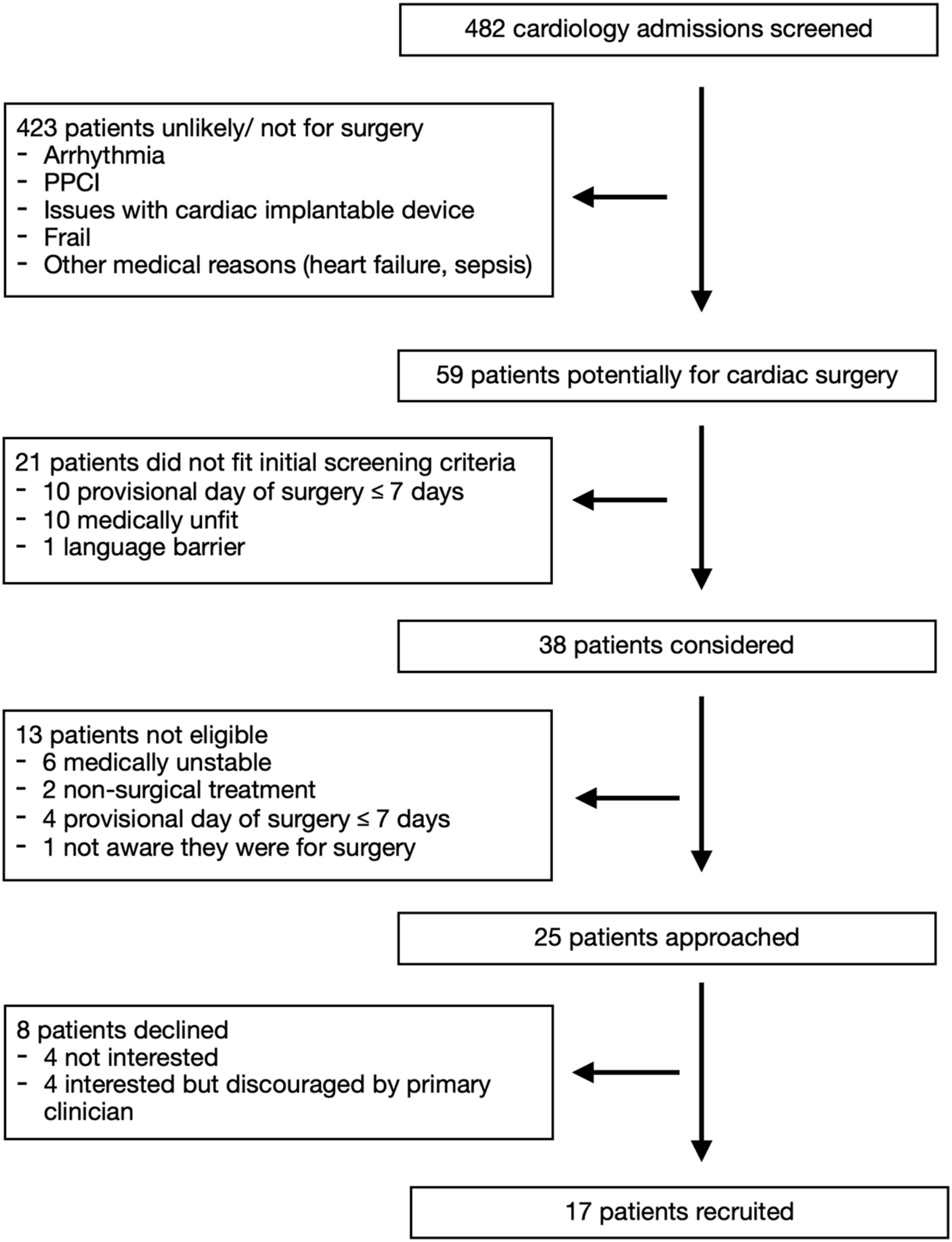
PPCI; Primary Percutaneous Coronary Intervention

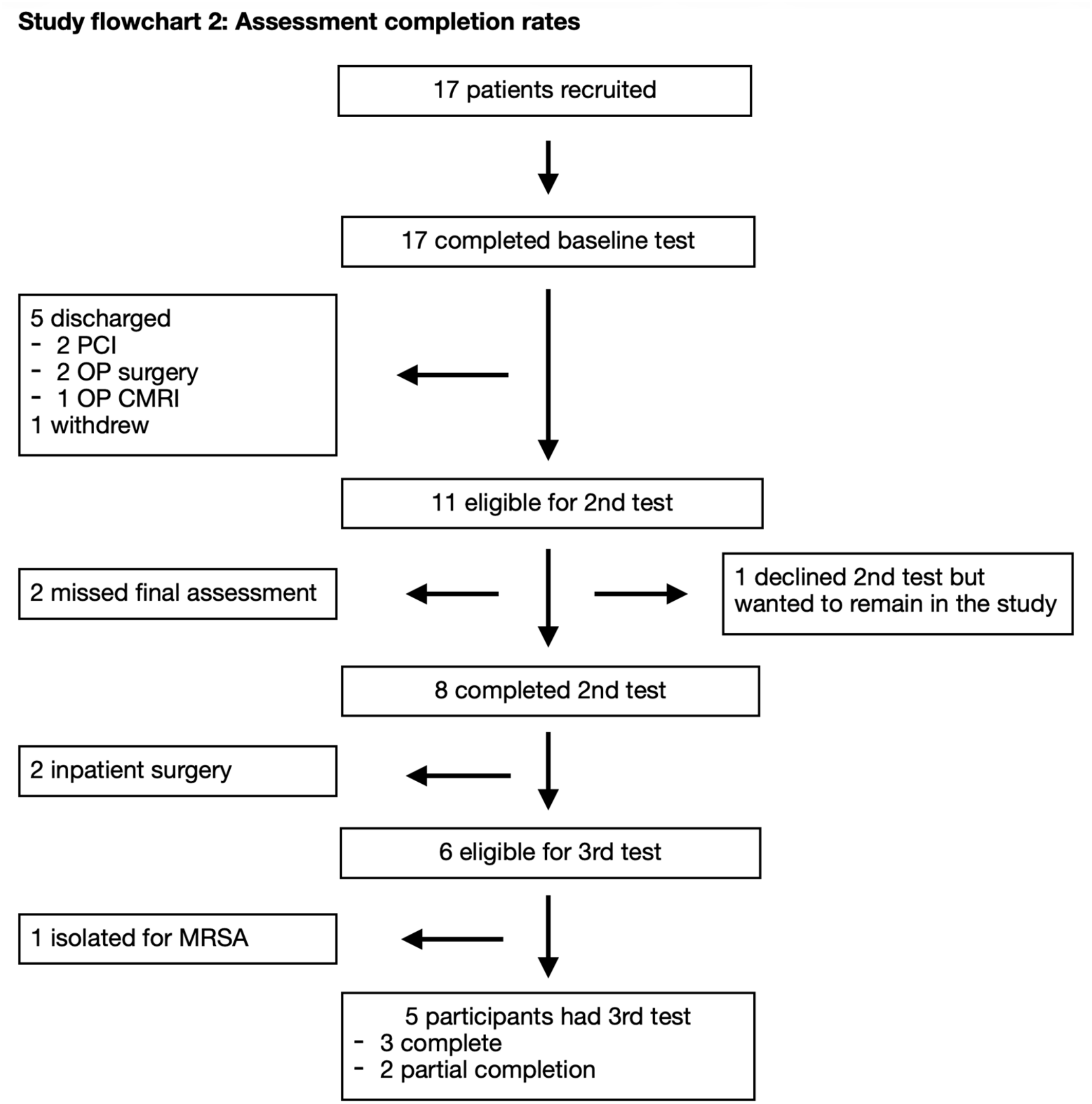
PCI; percutaneous Coronary Intervention, OP; outpatient, CMRI; cardiac magnetic resonance imaging, MRSA; Methicillin-resistant Staphylococcus Aureus

Participants who had chest pain at rest had EQ-5D-5L, CAQ, spirometer and HGT but not 6MWT. After the third test, the surgical team decided to send a further 2 patients home with an agreed date for surgery. It was unclear why this decision was taken. We termed this ‘outpatient surgery’ (OP surgery) as these were acute patients that presented from home for a planned surgery the day before the operation as opposed to acute patients that stay in hospital for ‘inpatient surgery’. Both patients completed the full test.

Table 1 shows a summary of the baseline characteristics and clinical data of the participants. A total of 17 patients were recruited. However, only 14 (82.4%) had cardiac surgery; 10 (71.4%) had inpatient surgery and 4 (28.6%) had outpatient surgery. The mean age of the recruits was 66.8 years (range 50-81) whereas the mean age for the inpatient surgical patients was 69.2 years (range 62-81). LV severity did not appear to determine likelihood of inpatient surgery. However, admitting CCS and NYHA scores may have an impact in deciding if the patient will have inpatient surgery. The most common operation performed was CABG. Participants spent a mean of 9.6 days (range 1-21 days) in the study. Participants waited a mean of 15.6 days (range 9-12 days) for their inpatient surgery and spent a further 7.7 days (5-16 days) after the surgery prior to discharge.

**Table 1:**
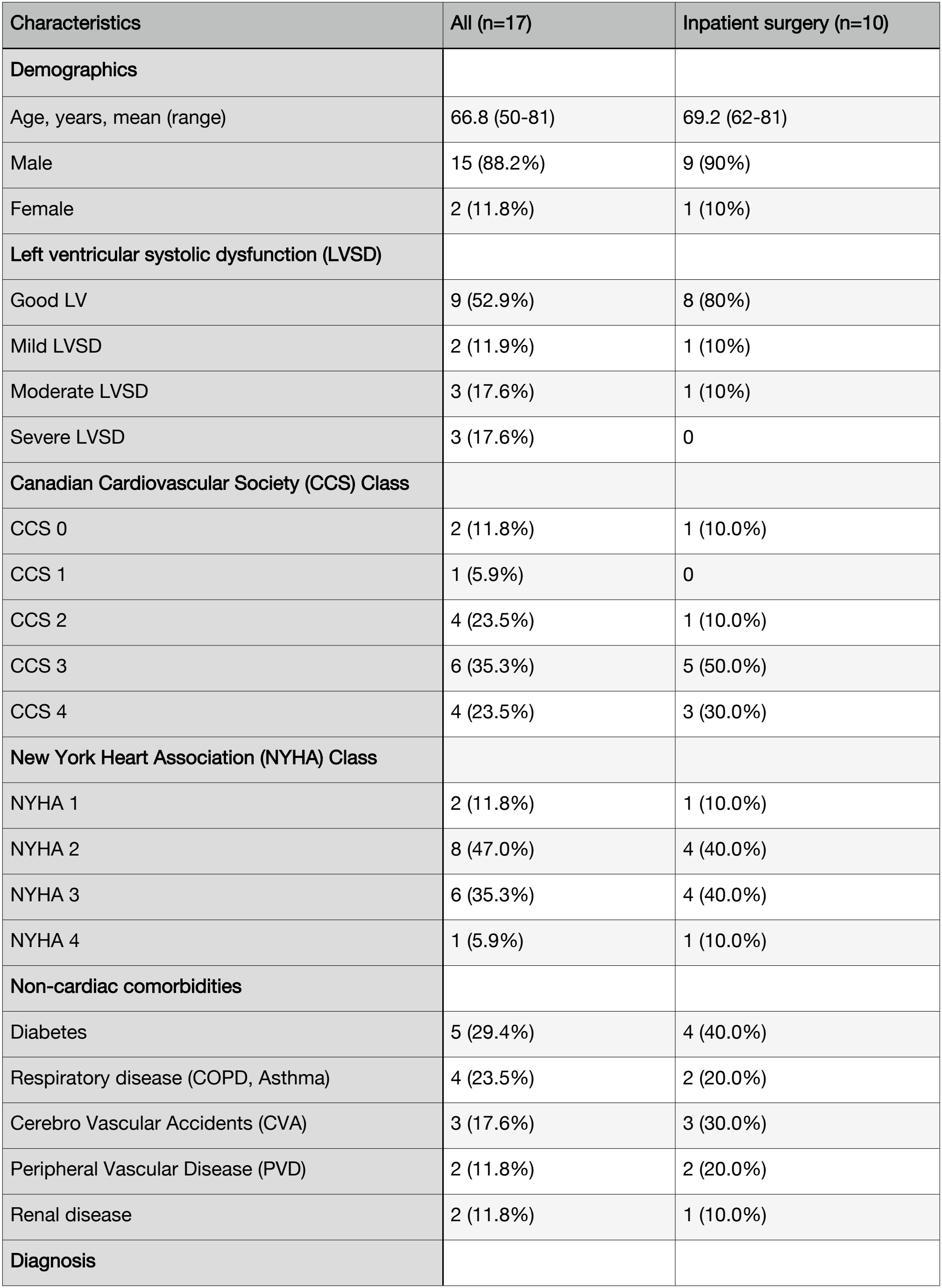
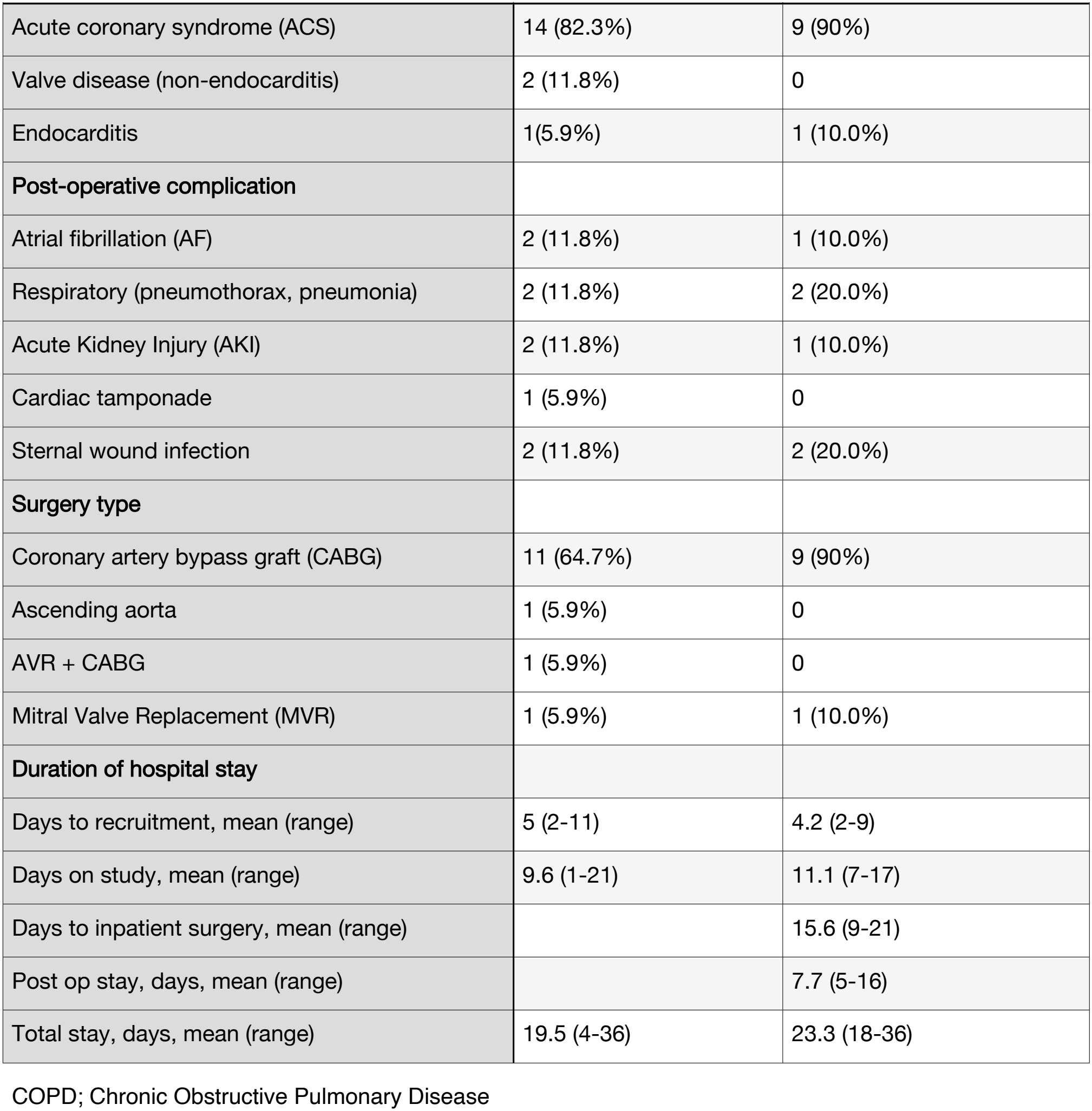
Baseline and clinical data.

### Intervention results

11 participants should have been offered exercise program as one participant withdrew and 5 were discharged within 5 days of recruitment. However, only 7 (63.6%) were seen by the physiotherapist and had subsequent exercise intervention due to staffing limitation. (Table 2)

**Table 2:** Aerobic exercise adherence and tolerability.

| Pt | No. session(s) offered | Adherence (%) | Tolerability (%) |  |  |  | Serious event(s) |
|---|---|---|---|---|---|---|---|
|  |  |  | 1 | 2 | 3 | x̄ |  |
| 1 | 1 | 100 | 26.7 | NA | NA | 26.7 | None |
| 2 | 3 | 100 | 40 | 100 | 100 | 80 | None |
| 3 | 2 | 100 | 100 | 100 | NA | 100 | None |
| 4 | 3 | 100 | 93.3 | 100 | 100 | 97.8 | None |
| 5 | 1 | 100 | 100 | NA | NA | 100 | None |
| 6 | 1 | 100 | 100 | NA | NA | 100 | None |
| 7 | 1 | 100 | 100 | NA | NA | 100 | None |

Pt; participant, x̄; mean

Adherence to the aerobic exercise was 100% for all 7 participants and there were no adverse events attributed to the aerobic exercise. (Table 2) Participant 1 only managed to exercise 26.7% of the allocated 15 minutes. This was because they had reached 50% HRR. Although they were completely asymptomatic and wanted to continue, we decide to stop the program and made adjustments for their next session. The participant then had PCI and was discharged before being seen for a second session of aerobic exercise. Unfortunately this meant that we could not assess how the changes in program would impact them.

Participant 2 managed 40% of the allocated 15 minutes of aerobic exercise due to feeling breathless and leg fatigue. (Table 2) They had past medical history of PVD, diabetes and obesity. Participant admitted to having a relatively sedentary lifestyle but was keen to improve their health. As such, they began walking around the ward daily. His exercise program was modified and the participant managed to complete 2 further exercise sessions without problems. That gave them a mean tolerability of 80% to the program.

Participant 4 had a one minute break during their first exercise session as they felt hot. After the break, the participant felt much better and wanted to continue with the exercise. The mean tolerability was 97.8%. (Table 2)

### Assessment Results

Table 3 shows the results of assessment performed on the 17 participants. The assessment were performed on day 0, 7 and 14 of the study. 2 participants did not perform 6 minute walk test on day 14 as they had recent chest pain at rest; 1 participant had aerobic exercise session one week prior to his day 14 assessment, the other did not receive any exercise intervention. The number of participants performing the test every week decreased as expected as they either had their surgery or were discharged as the week went on.

**Table 3:** Results of assessments.

| Assessment | Test 1 (Day 0) |  | Test 2 (Day 7) |  | Test 3 (Day 14) |  |
|---|---|---|---|---|---|---|
|  | N | Mean (SD) | N | Mean (SD) | N | Mean (SD) |
| **Hand grip test** | 17 |  | 8 |  | 5 |  |
| Left |  | 30.14 (9.48) |  | 32.19 (11.34) |  | 35.38 (5.86) |
| Right |  | 32.85 (10.12) |  | 34.95 (13.23) |  | 37.72 (8.37) |
| **Spirometry** | 17 |  | 8 |  | 5 |  |
| FEV1 |  | 2.28 (0.73) |  | 2.70 (0.75) |  | 2.64 (0.61) |
| FVC |  | 3.04 (0.73) |  | 3.36 (0.77) |  | 3.37 (0.50) |
| **6 minute walk test** | 17 | 365.57 (85.53) | 8 | 387.38 (87.34) | 3 | 414.00 (76.37) |
| **EQ5D5L** | 17 |  | 8 |  | 5 |  |
| Utility value | 16 | 0.86 (0.15) |  | 0.87 (0.11) |  | 0.83 (0.14) |
| VAS |  | 58.24 (20.91) |  | 65.13 (19.57) |  | 51.20 (24.39) |
| **CAQ** | 17 |  | 8 |  | 5 |  |
| Fear |  | 1.63 (0.78) |  | 1.59 (0.82) |  | 1.70 (0.19) |
| Avoidance |  | 1.37 (0.97) |  | 1.50 (0.92) |  | 1.36 (0.82) |
| Attention |  | 1.08 (0.60) |  | 1.10 (0.78) |  | 0.96 (0.48) |
| Overall |  | 1.40 (0.67) |  | 1.43 (0.72) |  | 1.40 (0.34) |

FEV<sup>1</sup>; Forced Expiratory Volume in 1 second, FVC; Forced Vital Capacity, VAS; visual analogue score, CAQ; Cardiac Anxiety Questionnaire

EQ5D5L utility value index score range from -0.285 (worse than dead) to 1 (perfect health),^(19)^ VAS range from 0-100. The EQ5D5L utility value on day 0 was based on 16 participants as one participant felt that they could not comment on domain 3; usual activity. Several participants mentioned an issue rating their ability to perform ‘usual activity’ as they were in hospital. However, most were satisfied to grade the question based on the ability to perform usual activities if they were at home.

CAQ is a 18 item questionnaire designed to a persons fear about heart sensations, avoidance of activities due to heart anxiety, and heart focused attention. ^(20)^ Each item is scored 0 (never) -4 (always) and the mean score for each sub-scale as well as overall score is shown above.

## Discussion

Prehabilitation in patients requiring inpatient cardiac surgery has never been done before. The protocol had to be modified during the study. Firstly identifying the best time to recruit to maximise the time spent in the study. Patients were initially recruited after the MDT decision but this reduced the time spent on the study. This study also found that although the MDT decision was initially for inpatient surgery, in some circumstances, this decision changed weeks down the line; not because the patients become less acute, but they were staying in hospital and contracting infections. SR tried recruiting before the MDT based on the clinical team’s opinion on best treatment for the patient. This led to several discharges after the MDT meeting.

In the original protocol, we aimed to repeat the final test the day before surgery. However, this quickly proved problematic as the date of surgery could change at short notice. As such, the test were fixed on day 7 and 14. The aerobic exercise component was bespoke and fully monitored by a physiotherapist. This allowed adjustments to suit the individual. Some participants had a sedentary lifestyle which impacts their ability to exercise. The exercise target was moderate exercise intensity which correspond to 40-70% heart rate reserve (HRR) and Borg Rating of Perceived Exertion (RPE) of 10-14. At the start of the program, we aimed for exercise up to 50% HRR. However, the team soon found that participants were exceeding the HRR within minutes but the RPE was below 10 and participants were asymptomatic. Following the team discussion, it was agreed that we will aim for 70% HRR; this change did not appear to impact symptoms but allowed participants to achieve more during their aerobic exercise.

Once eligible participants were identified, 84% participants approached were interested. However, after discussion with their primary clinicians, 4 participants decided not to participate. It is unclear why they were discouraged. We recruited 17 participants which gives a recruitment rate of 1.5 participants per month. Clinician buy-in is essential to improve recruitment rate. Only one participant withdrew from the study giving a retention rate of (94.1%). The completion rate for assessments throughout the trial was above 70%. Whilst tolerance, compliance and adherence to the program was good, our study highlighted the importance of staffing as a barrier to delivery; only 63.6% were seen by the physiotherapist and received exercise programs. There were no adverse events or safety concerns reported with regards to inpatient prehabilitation during our study.

We were unable to comment on the impact of the interventions on the secondary outcome. First, it is expected that the number of patients remaining in the study reduces each day as they undergo surgery. Only 50% participants had repeat assessments.

Secondly, 7 participants received exercise intervention. These numbers are too small to identify any changes in the assessment results due to the interventions. The attrition rate leads to loss of statistical power and attrition bias which meant that we were unable to perform inferential statistics.

Enhanced recovery after cardiac surgery (ERAS-cardiac) society recommends prehabilitation to include exercise, psychological and nutrition component.^(21)^ Our study included exercise and psychological components.^(18)^ The nutrition component was intentionally omitted after our PPI work revealed that participants did not want education on nutrition and the hospital dietician discouraged significant dietary changes or supplements due to the short time line.^(18)^

## Limitations

At the time of conception, there were no trials on prehabilitation in acute inpatients awaiting cardiac surgery. This is a single-arm, single-centre feasibility trial that recruited 17 participants; 7 receiving the intervention. As such it is underpowered to assess for safety and efficacy of the intervention or assessments.

The study was limited by the lack of funding. The study did not have a physiotherapist or psychotherapist dedicated for this study. This meant that there were 10 ‘missed opportunities’ for exercise intervention. However, this would be reflective of the outcome if prehabilitation was delivered within the current healthcare system.

The study was conducted in a tertiary centre. However, many patients have spent a period of time in a DGH. As confidence in acute prehabilitation increases, the program could be started in the DGH. This will increase the duration of time for prehabilitation without changing the actual course of definitive treatment.

## Conclusion

Measures of feasibility includes recruitment, retention, implementation, acceptability, tolerability and adherence towards the intervention. Prehabilitation in acute inpatients for cardiac surgery is a novel idea that appears well received, tolerated and adhered to by patients. Unfortunately recruitment, and implementation of the intervention has been limited by the lack of funding and reluctance from the clinician. Future trials without adequate funding are unlikely to succeed. By the same token, success of a well funded trial is unlikely to be translatable in real clinical practice without equivalent budget. Some clinicians appear hesitant to embrace prehabilitation. There is no work currently done to ascertain clinician opinion or address their reluctance on prehabilitation.

## Data Availability

The data underlying this article will be shared on reasonable request to the corresponding author.

## Authors’ contributions

Conceptualisation: SR, LI

Methodology: SR, AH, LI

Supervisors: DY, GD, LI

Writing original draft: SR

Writing- review and editing: all authors

## Acknowledgements

We would like to thank all of the co-investigators involved in making this trial possible. Lili Dixon (Clinical psychologist) and Francesca Leoni (Cardiac Surgical registrar) for their assistance in developing the psychoeducation booklet, Jessica Prance (Physiotherapist) for their assistance in providing aerobic exercise. Last but not least, we would like to acknowledge the contribution of Professor Dumbor Ngaage who has contributed to the protocol and providing advice throughout the study but has unfortunately passed on in March 2026.

## Data availability statement

The data underlying this article will be shared on reasonable request to the corresponding author.

## Appendix

CONSORT 2010 checklist of information to included when reporting pilot or feasibility trial

